# POOLING FOR SARS-COV-2 CONTROL IN CARE INSTITUTIONS

**DOI:** 10.1101/2020.05.30.20108597

**Authors:** Jorge J Cabrera, Sonia Rey, Sonia Pérez, Lucía Martínez-Lamas, Olaia Cores-Calvo, Julio Torres, Jacobo Porteiro, Julio García-Comesaña, Benito Regueiro

**Affiliations:** Microbiology and Infectology Research Group, Galicia Sur Health Research Institute (IIS Galicia Sur). SERGAS-UVIGO.; Microbiology Department, Complexo Hospitalario Universitario de Vigo (CHUVI), Sergas. Vigo, Spain.; CINTECX, Universidade de Vigo, GTE. Vigo, Spain; Management Department. Complexo Hospitalario Universitario de Vigo (CHUVI), Sergas. Vigo, Spain.; Microbiology and Parasitology Department. Medicine and Odontology. Universidade de Santiago. Santiago de Compostela, Spain.

**Keywords:** SARS-CoV-2, pooling, detection, care homes, disability institutions, low prevalence

## Abstract

Workers and residents in Care Homes are considered at special risk for the acquisition of SARS-CoV-2 infection, due to the infectivity and high mortality rate in the case of residents, compared to other containment areas. The aims of the present study, based in our local experience, were (a) to describe SARS-CoV-2 prevalence in institutionalized people in Galicia (Spain) during the Coronavirus pandemic and (b) to evaluate the expected performance of a pooling strategy using RT-PCR for the next rounds of screening of institutionalized people.

Distribution of SARS-CoV-2 infection at Care Houses was uneven. As the virus circulation global rate was low in our area, the number of people at risk of acquiring the infection continues to be very high. In this work, we have successfully demonstrated that pooling of different groups of samples at low prevalence clusters, can be done with a small average delay on quantification cycle (Cq) values. A new surveillance system with guaranteed protection is required for small clusters, previously covered with individual testing. Our proposal for Care Houses, once prevalence zero is achieved, would include successive rounds of testing using a pooling solution for transmission control preserving testing resources. Scale-up of this method may be of utility to confront larger clusters to avoid the viral circulation and keeping them operative.

## INTRODUCTION

Severe acute respiratory syndrome coronavirus 2 (SARS-CoV-2), the causal agent of Coronavirus disease 2019 (COVID-19), emerged in late 2019 in China. More than 5,016,000 infections and more than 328,000 deaths have been attributed globally [1]. As people over 70 years old are particularly likely to develop a severe infection [2,3], surveillance of Care Homes has been critical to limit the mortality rate. In Galicia (Spain), the local health authority (Servizo Galego de Saude, SERGAS) established policies of self-isolation and quarantine in conjunction with Public Health to prevent SARS-CoV-2 from spreading. However, they also focused their efforts in Care Homes from an early stage. Public health interventions included testing people institutionalized in Care Homes and all related workers. These measures allowed the preservation and functionality of these facilities.

Direct viral detection is useful to identify people with potential SARS-CoV-2 transmission risk. Real time RT-PCR is the most common method used for viral RNA detection in the nasopharyngeal swab, but limited stocks and restrictions in test capacity prevented a higher number of PCR tests per day. Identification and isolation of PCR positive infected people and contact tracing had been the main strategy followed to limit virus spreading. However, considering that less than 5% of the Spanish population is estimated to have been infected, lifting lock-down measures will also require the introduction of new care strategies of surveillance relevant for institutionalized people and other significant clusters.

Pooling strategies have proven to preserve SARS-CoV-2 testing resources. According to recent publications, group testing could save reagents and time with an increase in testing capability of the 69% for an incidence rate of SARS-CoV-2 infection of 10% or less [4] [5] [6] [7]. Pooling could be associated with a decrease in detection related to the increase of the limit of detection of the individual sample [8] [9]. Main limitations could be the pooling preanalytical step, the sample viral load or the global sensitivity of the diagnostic procedure [10].

The rationale in this study is to develop a new strategy based on initial individual identification of positive coronavirus cases in order to organize low prevalence clusters, followed by a serial pooling strategy testing of these clusters, in order to control areas free of virus circulation, allowing them to be fully operative.

## MATERIALS AND METHODS

### 1. Samples

Nasopharyngeal swab samples were obtained from residents and workers at Care Homes in Galicia (March to May 2020). Several viral transport mediums were used, mainly Vircell Transport Medium (Vircell, Granada, (Spain)) and *δ* swab® Transport Medium (Deltalab, Rubí, (Spain)). Samples were processed on the first 48 hours and frozen for storage on the first 72 hours. When two samples were received from the same patient (follow-up), only the first sample was considered for the prevalence study. The study was performed in accordance with protocol (2020/298), approved by the Galician network of committees of research ethics.

### 2. SARS-CoV-2 testing

#### 2.a) Care Homes screening

Screening was performed with the cobas® SARS-CoV-2 test (Roche Diagnostics, NJ, USA) on the cobas® 6800 system (Roche Diagnostics). Samples were mixed 1:1 with cobas® omni lysis reagent (43% guanidine thiocyanate) and incubated for 15 min before loading onto the cobas® 6800 system for viral inactivation. No further manual steps were required during the workflow. This test includes fully automated sample preparation (extraction and purification of 400 μl of sample and added internal control RNA (RNA IC) molecules) followed by PCR amplification and detection. Automated data management is performed by the cobas® 6800 software, which assigns test results for all tests. External controls (positive and negative) are processed in the same way within each run.

Two targets were amplified in the multiplex one-step rRT-PCR: The ORF1/b non-structural region that is unique to SARS-CoV-2, and a conserved region in the structural protein envelope E-gene for pan-Sarbecovirus detection.

A sample was positive if at least ORF1/b was detected. A sample was also considered positive if only E-gene was detected twice. A sample was negative if only IC is detected. Any Cq below cycle 40 was considered as positive.

#### 2. b) Analytical sensitivity

Cobas® SARS-CoV-2 test, Allplex™2019-nCoV (MagCore® HF16 Plus system extraction) and Allplex™2019-nCoV (STARlet) assays were run in triplicate using 2 × 10^4^ DNA copies/mL and 2-serial dilutions of SARS-CoV-2 standard at starting concentrations of 2 × 10^3^ DNA copies/mL to 31.25 DNA copies/ml. Additional dilutions were tested: 40, 20 and 2 DNA copies/ml. The sensitivity of each assay was considered as the lowest concentration of SARS-CoV-2 giving a positive result for any target.

The EDX SARS-CoV-2 Standard (Exact Diagnostics, TX, USA) containing 200,000 copies/mL of synthetic RNA transcripts from five gene targets (E, N, ORF1ab, RdRP and S Genes of SARS-CoV-2) was used.

#### 2.c) Pooling

Pooling was performed by the QIAgility instrument (QIAgen) designed to perform automated PCR set-up and liquid handling actions in molecular biology applications. A volume of 50-150 μL of each sample was included in the pool.

#### 2.d) Pool positivity assessment

Pools of negative samples and 1 positive sample and the corresponding positive sample were processed using the MagCore® HF16 Plus system (RBC Bioscience) and the Allplex™2019-nCoV assay (Seegene In, Seoul, South Korea). The automated workstation MagCore was used for acid nucleic extraction (400 μL of sample and added RNA IC). Allplex™2019-nCoV assay was used for the amplification step on the CFX-96 system (Bio-Rad Laboratories, Hercules, CA, USA) after manual PCR set-up. Unextracted positive control was added during the PCR set-up.

Three viral targets were detected: RNA-dependent RNA polymerase (RdRP) and nucleocapsid (N) genes specific for SARS-CoV-2, and a conserved region in the E gene. Analysis of results was done using specific 2019-nCoV Seegene viewer software. A sample was considered positive if at least one target was detected.

#### 2.e) Pooling proof of concept

Two simulations were tested by pooling. Sample distribution within pools was randomly performed. Results of samples individually tested by cobas® SARS-CoV-2 test were compared with results of samples tested in pools by the STARlet instrument (Microlab) with STARMag 96 × 4 Universal Cartridge Kit for automated extraction (200 μL of sample and added RNA IC) and the Allplex™2019-nCoV Assay PCR set-up.

### 3. Statistical analysis

Prevalence and distribution of positive detection in Care Homes was calculated. Medium, mean, range and quartiles were calculated for Cq values of each target detected by cobas®6800 from screening of Care Homes. Mean and range were calculated for each target Cq value and for each target Cq value difference between individual or pooled samples (R version 3.5.1 http://www.R-project.org/). The Cq values were considered as 41 in case of undetectable result.

## RESULTS

### SARS-CoV-2 prevalence in institutionalized people in Galicia (Spain) during the Coronavirus pandemic

During the Coronavirus pandemic, SARS-CoV-2 prevalence was calculated for 25,386 people from 306 Galician Care Homes: 16477 residents, 8,599 workers and 310 not specified. The mean age of workers and residents was 44.25 years (min 18, max 69) and 80.07 years (min 3, max 109), respectively. SARS-CoV-2 was detected in 852 people (3.36%). The distribution of institutions by SARS-CoV-2 prevalence is shown in Table 1 and figure 1. A total of 263 institutions (19,091 people) had SARS-CoV-2 prevalence zero. A total of 282 institutions (21,861 people) had SARS-CoV-2 prevalence <5%.

**Table 1.** Distribution of care institutions by SARS-CoV-2 prevalence

| SARS-CoV-2 prevalence (%) | Care Institutions (n) | People (n) |
| --- | --- | --- |
| 0 | 263 | 19,091 |
| >0-1 | 8 | 1,783 |
| >1-2 | 8 | 866 |
| >2-3 | 1 | 43 |
| >3-4 | 2 | 78 |
| >4-5 | 0 | 0 |
| >5-6 | 1 | 319 |
| >9-10 | 1 | 80 |
| >10-20 | 11 | 1,817 |
| >20-30 | 1 | 77 |
| >30-40 | 4 | 549 |
| >40-50 | 2 | 320 |
| >50-60 | 4 | 363 |
| **Total** | **306** | **25,386** |

**Figure 1.**
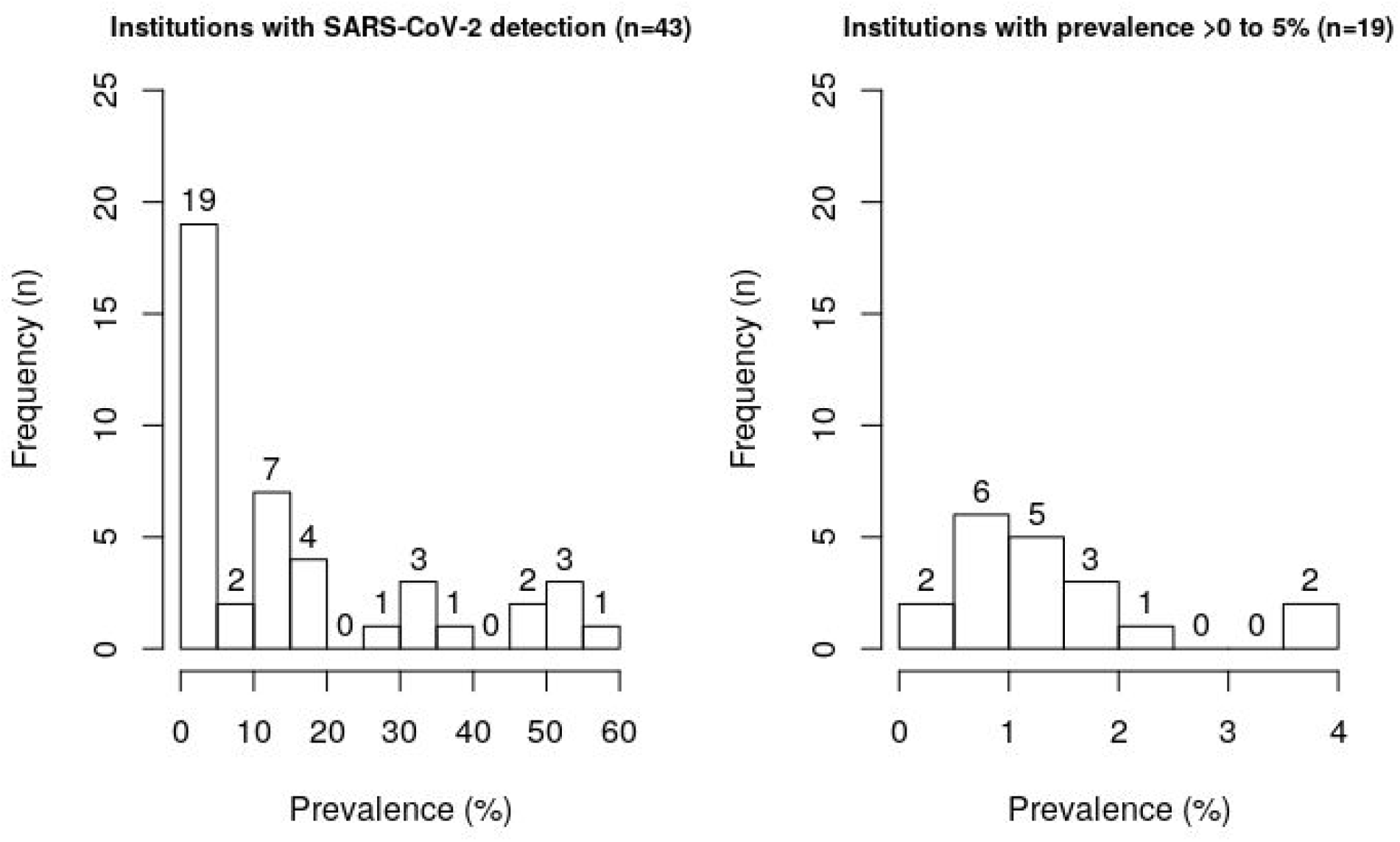
Distribution of Care Institutions by SARS-CoV-2 prevalence, excluding those with prevalence zero

The cobas® SARS-CoV-2 test yielded a positive result in 803 cases (94.2%, both targets positive). Isolated ORF1/b was detected in 9 cases (1.1%). Isolated E gene was detected in 40 cases (4.7%). Cq value distribution for positive samples is shown in table 2 and figure 2 for both cobas® SARS-CoV-2 targets. For 90% of positive samples, ORF1b Cq value was lower than 32.9 and E gene Cq value was lower than 35.9.

**Table 2.** Screening of care institutions. Quantification cycle distribution of positive samples detected by cobas® SARS-CoV-2 test.

| Target | minimum | 1st quartile | medium | mean | 3rd quartile | maximum | Indetectable target (n) |
| --- | --- | --- | --- | --- | --- | --- | --- |
| ORF1b | 15.03 | 21.86 | 26.41 | 26.40 | 31.36 | 35.86 | 69 |
| Gen E | 15.41 | 22.55 | 27.54 | 27.89 | 33.60 | 39.06 | 9 |

**Figure 2.**
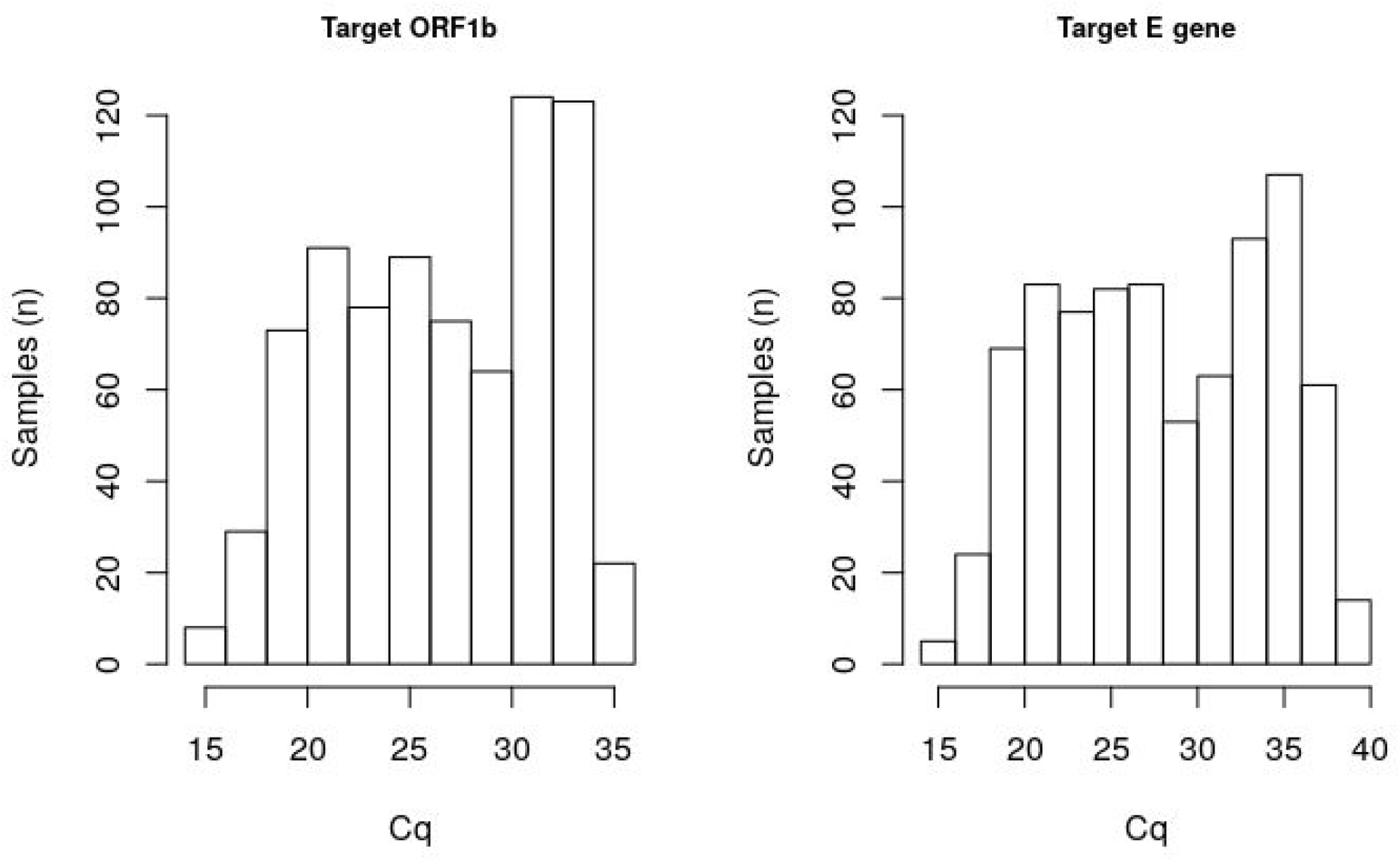
Distribution of quantification cycle (Cq) values for each target of cobas® SARS-CoV-2 test (ORF1/b and E gene).

### SARS-CoV-2 assays limit of detection

Sensitivity was 31.25 copies/μl (6.75 copies/reaction) for cobas® SARS-CoV-2 test on the cobas® 6800 system, 125 copies/μl (6.67 copies/reaction) for Allplex™2019-nCoV assay after nucleic acid extraction with MagCore® HF16 Plus system and 250 copies/ml (4 copies/reaction) for Allplex™2019-nCoV assay after nucleic acid extraction with STARlet system (Hamilton (USA)).

### Pool positivity assessment

According to other authors calculations (https://www.chrisbilder.com/shiny), for prevalence between 1 and 2%, sensitivity 95% and specificity 100%, the optimal pool size would be between 25 and 16 samples and the optimal sub pool size would be between 4-5 samples. In order to minimize the false negative factor for pooled testing recently defined (Hanel and Thurner 2020) and to standardize the pooling method, pools of 20 samples (P20) and sub pools of 5 samples (SP5) were selected.

Test performance of 26 P20 s and 14 SP5 was studied. Positive samples were selected between those originally tested by cobas® SARS-CoV-2. Each pool was made up of negative samples and one positive sample with Cq values for the two cobas® targets within the first, second or third quantile of total Cq values (mean 27.43 and 28.68 for ORF1/b and E gene, respectively). All positive samples individually detected by cobas® SARS-CoV-2 test yielded positive results when tested in pools of 20 or 5 samples by Allplex™2019-nCoV assay. Mean and range of Cq values for E gene, RdRp and N gene for individual samples and their differences in the corresponding pools are shown in table 3. Mean delay in the Cq values (Cq pool–Cq positive sample) was 5.02 cycles for the P20 and 2.85 cycles for the SP5. The N gene was not detected by Allplex™2019-nCoV assay in one specific sample independently of pooling or individual testing.

**Table 3.**
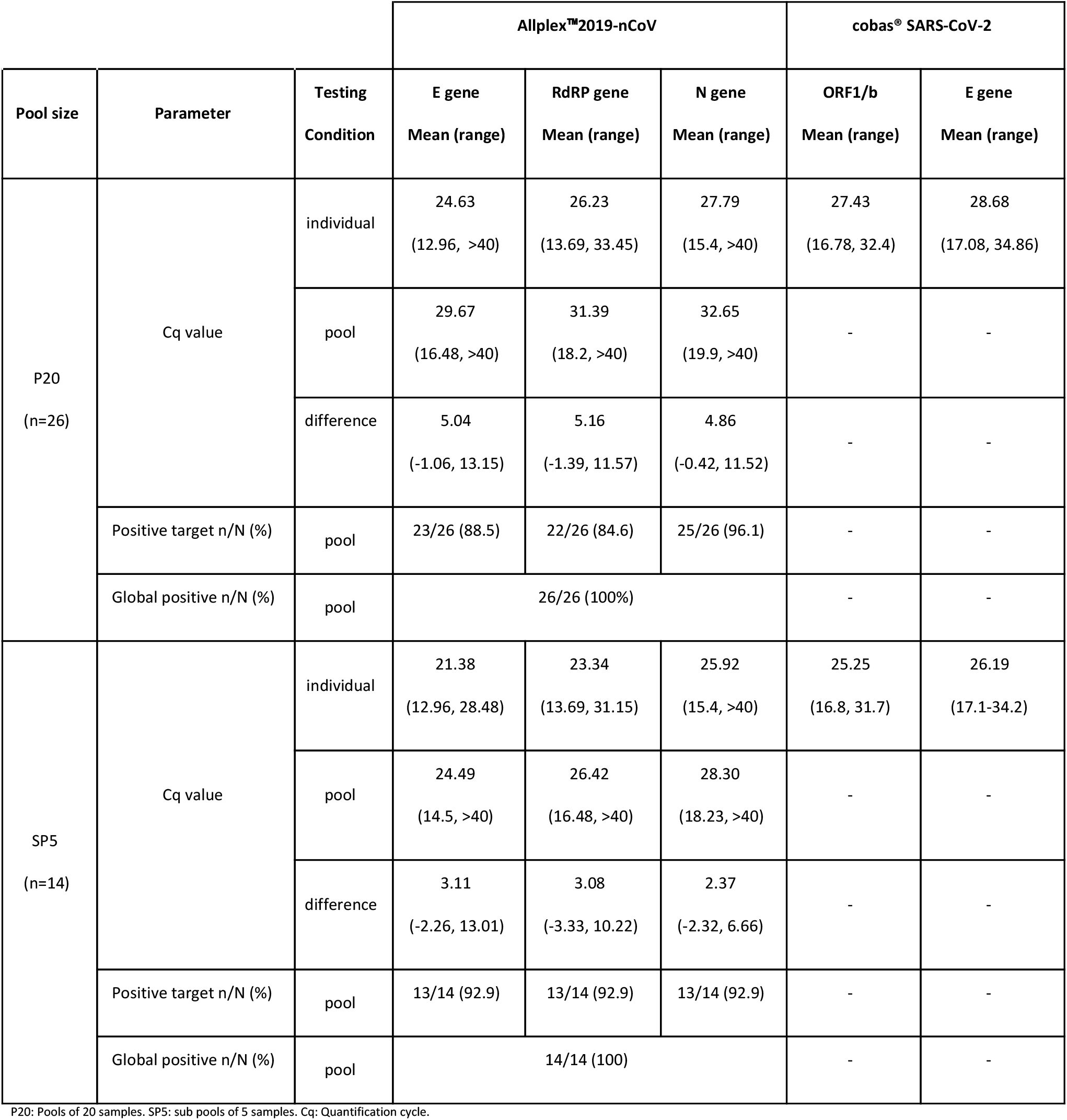
Differences in quantification cycle (Cq) values between pool and individual testing (mean and range) in Allplex™2019-nCoV assay:

| Pool size | Parameter | Testing Condition | Allplex™2019-nCoV |  |  | cobas® SARS-CoV-2 |  |
| --- | --- | --- | --- | --- | --- | --- | --- |
|  |  |  | E gene<br>Mean (range) | RdRP gene<br>Mean (range) | N gene<br>Mean (range) | ORF1/b<br>Mean (range) | E gene<br>Mean (range) |
| P20<br>(n=26) | Cq value | individual | 24.63<br>(12.96, >40) | 26.23<br>(13.69, 33.45) | 27.79<br>(15.4, >40) | 27.43<br>(16.78, 32.4) | 28.68<br>(17.08, 34.86) |
|  |  | pool | 29.67<br>(16.48, >40) | 31.39<br>(18.2, >40) | 32.65<br>(19.9, >40) | - | - |
|  |  | difference | 5.04<br>(-1.06, 13.15) | 5.16<br>(-1.39, 11.57) | 4.86<br>(-0.42, 11.52) | - | - |
|  | Positive target n/N (%) | pool | 23/26 (88.5) | 22/26 (84.6) | 25/26 (96.1) | - | - |
|  | Global positive n/N (%) | pool | 26/26 (100%) |  |  | - | - |
| SP5<br>(n=14) | Cq value | individual | 21.38<br>(12.96, 28.48) | 23.34<br>(13.69, 31.15) | 25.92<br>(15.4, >40) | 25.25<br>(16.8, 31.7) | 26.19<br>(17.1-34.2) |
|  |  | pool | 24.49<br>(14.5, >40) | 26.42<br>(16.48, >40) | 28.30<br>(18.23, >40) | - | - |
|  |  | difference | 3.11<br>(-2.26, 13.01) | 3.08<br>(-3.33, 10.22) | 2.37<br>(-2.32, 6.66) | - | - |
|  | Positive target n/N (%) | pool | 13/14 (92.9) | 13/14 (92.9) | 13/14 (92.9) | - | - |
|  | Global positive n/N (%) | pool | 14/14 (100) |  |  | - | - |

P20: Pools of 20 samples. SP5: sub pools of 5 samples. Cq: Quantification cycle.

### Proof of concept

Two simulations were retrospectively tested in pools with Allplex™2019-nCoV assay (STARlet) using the following algorithm: P20, SP5 when positive, individual analysis when positive.

A first simulation was performed with 100 samples with 2% prevalence. Five P20 were tested. As 2 positive pools were obtained, 8 SP5 were processed. Two SP5 were positive, so 10 samples were tested individually. Two samples were positive. Number of tests was reduced 77% (0.23 tests per individual). Strategy and Cq was shown in figure 3.

A second simulation included 60 samples with 1.7% prevalence. Three P20 were tested. One P20 was positive, so 4 SP5 were tested. One SP5 was positive so 5 samples were tested individually. One sample was positive. Number of tests was reduced by 80% (0.20 tests per individual).

**Figure 3.**
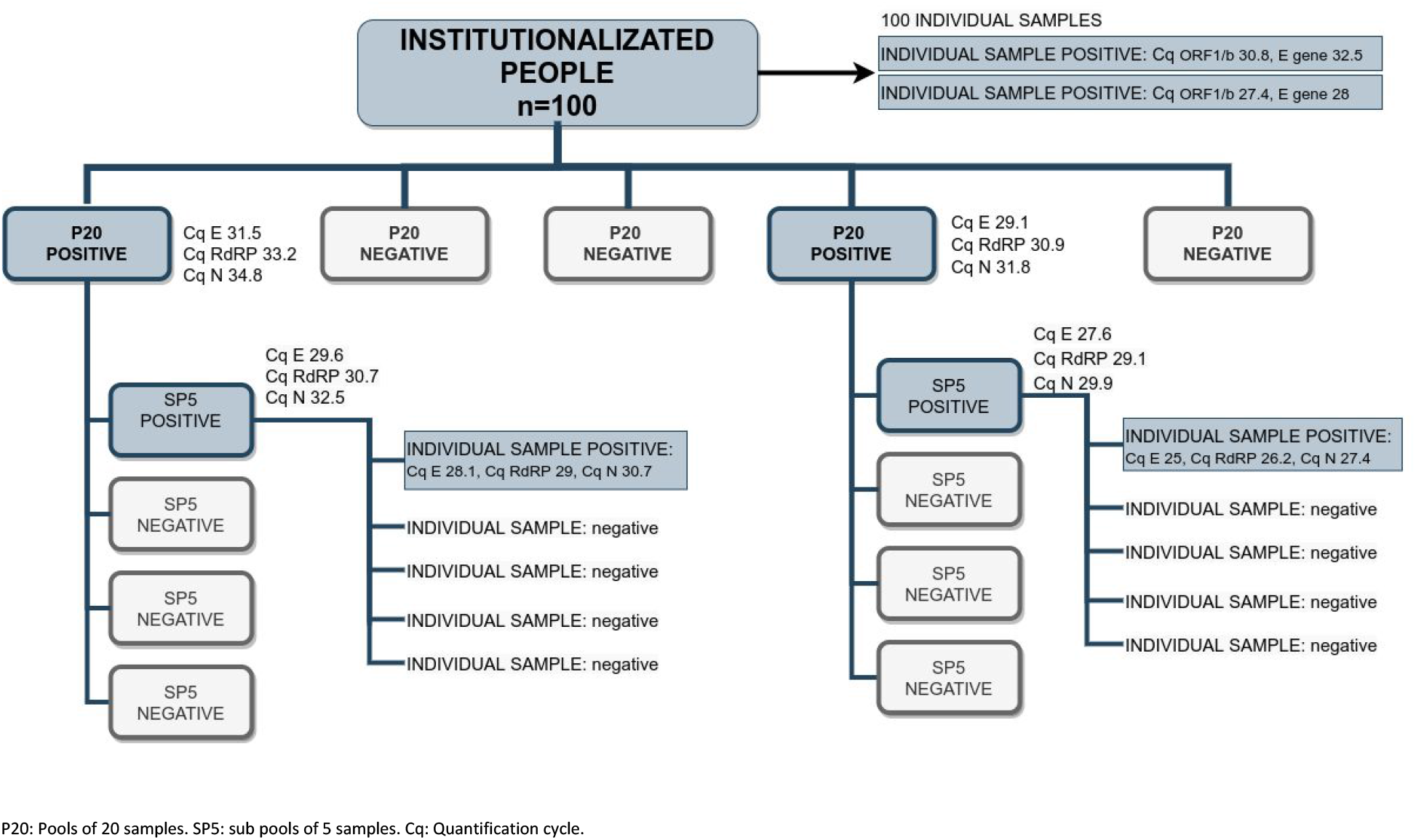
Pooling testing of 100 samples from Care Institutions with 2% prevalence. P20: Pools of 20 samples. SP5: sub pools of 5 samples. Cq: Quantification cycle.

## DISCUSSION

There are large ongoing seroprevalence studies in Spain. Preliminary results of the first round have shown a global prevalence of 5% in Spain, varying from 1.1% (Ceuta) to 11.3% (Madrid) [11]. When definitive results are available, control strategies will be adapted to local situations. The results of smaller studies showed a seroprevalence of 11.2% in a high-risk population to acquire SARS-CoV-2 infection (578 Health care workers) in Barcelona [12]. For Galicia, preliminary results from the general population showed a seroprevalence between 1.15% (22,471 people) [13] to 2.1% [11]. All these seroprevalence data and global viral prevalence around 3% at Care Houses in Galicia (Spain), reported in the present study, suggest that the virus circulation rate has been low in our area. Therefore, the number of people at risk of acquiring the infection continue to be very high. In the current de-escalation stage, different COVID-19 containment strategies will be needed. Previous studies have studied how the SARS-CoV-2 spreads from person to person and the contribution of different sources of virus infection. The role of presymptomatic people: It has been estimated that up to 47% of infections happen from presymptomatic individuals, 40% from symptomatic individuals, 10% from the environment and 6% from asymptomatic people who never show symptoms [14] in line with others studies [15] [16]. To control the spread of the virus, it is essential to detect as many infected individuals as possible, as quickly as possible to trace down and test possible contacts [17]. To this end, it is important to detect presymptomatic patients and increase the number of people tested. In this context, sample pooling strategies can substantially increase the diagnostic capacity of laboratories and help to improve infection control.

The initial approach to this pandemic was individually tested by RT-PCR for the presence of SARS-CoV-2. Using this strategy we performed the screening of 306 Care Homes and a total of 25,386 determinations in workers and residents. Contact trace was performed and infected people were isolated. The need for a systematic control of the safety of these clusters, which had the highest mortality rate, led us to develop an alternative strategy. We proposed the use of pooling strategies, conventionally used to preserve SARS-CoV2 testing, to test areas free of virus circulation. Our results showed a prevalence <2% for more than 85% people in Care Houses. In these settings, pooling could achieve maximum usefulness. This prevalence data was used to search the optimal configuration of pools and sub pools. After reviewing the literature, and due to the absence of accumulated experience with this type of strategy for SARS-CoV-2, a more conservative size pool of 20 and sub pool of 5 samples was chosen [4] [5] [18].

Two tests authorized by Food and Drug Administration (FDA) for emergency use were available at our laboratory: The Roche Molecular Systems, cobas® SARS-CoV-2 test for use on the cobas® 6800 system and the Seegene, Allplex™2019-nCoV Assay. They were authorized for individual qualitative detection of nucleic acid from SARS-CoV-2 in several respiratory samples from people who were suspected to have COVID-19 by their health care provider. Both tests have shown suitable specificity and sensitivity for clinical diagnosis, but specific studies will be required for assessing their performance in pooling conditions. The choice of Seegene for pooling processing was due to the flexibility and adaptability in the automation process available in our laboratory. The working system as a whole is an open system with each individual step (pipetting, nucleic acid extraction and amplification) with modifying possibilities to find the most appropriate extraction conditions or interpretation could be adapted for the process optimization. Roche is a fully automated system where extraction, amplification or interpretation conditions cannot be modified by the user. Additionally, the possibility of detecting three targets instead of two, could increase the possibilities of detection in case of unknown mutations. Although it has been established a moderate mutation rate of SARS-CoV-2 [19] [20] [21] in comparison with other ARN viruses, some variants might compromise the detection [22] [20]. This could be critical in the case of the E gene, which has been found as the only target in a high proportion of samples with cobas® SARS-CoV-2 test in our population.

All positive samples when tested individually by cobas® SARS-CoV-2 test were positive in P20 and SP5 by Allplex™2019-nCoV assay. As previous studies [18] [8] [9], our results using pools showed an increase in the Cq value between pooled tests and individual positive samples. The average increase for P20 was of five cycles whereas P5 showed an average increase of three cycles. This increase in Cq has not carried out loss of sensitivity in pools of five or twenty samples with Cq value within the first three quartiles observed in our population.

### Proposed methodology

Here there is our proposal for introducing the pooling strategy in Care Institutions: When an institution with prevalence zero is characterized, successive rounds of testing would be the option for transmission control. The maximum interval between rounds would be adjusted to avoid the loss of detection of infected people who could be in a phase of low viral load. The incubation period has been reported to be highly variable with an estimated average of 5-6 days [23] [24] [25]. Viral load could be detectable 2-3 days before onset for a median of 20 days after symptoms onset, although levels drop significantly after day 10 [26] [16]. In this study we have focused on demonstrating that any pool containing individual samples from highly infectious people would be detected. Cq values associated with low viral load were excluded (fourth quartile) as this would not be the expected situation. In fact, the expected arrival of new cases would be of presymptomatic or initial symptomatic contagious individuals with viral loads reaching their peak, as multiple rounds of screening are being considered and positive and symptomatic people are excluded from pooling testing. In this situation, considering a doubling time higher than 14 days, test rounds of 14 days will allow us to detect any new highly infectious person. This would allow us to introduce highly effective contact tracing and case isolation. This strategy would be enough to control new outbreaks of COVID-19. The frequency of rounds should also be adjusted by local doubling time and serial interval in the case that a positive appears in the cluster. It seems reasonable to adapt time for new testing according to virus circulation, local rate of infection or risk severity and population tolerance of the sampling method.

Pooling strategies could be extended to other settings allowing large scale screening. Different conditions could influence size, distribution, and number of clusters to test. These clusters would be associated with different optimal pool sizes or optimal frequency of testing cycles. The development of specific pooling strategies will be necessary for the process optimization. It will even require the collaboration of Human resources in many settings. It would be very interesting to study the impact of including new parameters as self-sampling, health auto-checking or individual immune state in the pooling assignment by clusters.

Limitations of this study were the limited number of samples included. Testing more negative samples would be desirable to assess specificity and the risk of contamination along the processing. Cq values were not directly comparable because of differences in sample pre processing step (dilution) and the retrospective design of the pooling analysis.

This work has shown the prevalence of SARS-CoV-2 in Spanish Care Homes during the Coronavirus pandemic. Prevalence differences shown between Institutions should address the interventions for viral transmission control. Few studies have assessed the performance of pooling for SARS-CoV-2 detection by rRT-PCR in real conditions, especially when aiming to keep areas free of virus circulation to be operative and functional. Sample pooling seems to be a new testing strategy relevant for maintaining low level or no transmission among institutionalized people. Further studies with self-sampling methods, modular systems and more specific pooling strategies will be necessary for the process improvement.

## Data Availability

The authors confirm that the data supporting the findings of this study are available within the article.

## ACKNOWLEDGEMENTS

To the Servizo Galego de Saúde, Consellería de Sanidade and Xunta de Galicia for supporting this study. To Dr Vázquez Almuiña for his suggestions. To all the professionals who collaborated in information management, sampling, supplies, contact tracing and organization at all levels. Especially to the technician staff of the Microbiology Department of the CHUVI. To Laura Regueiro for her assistance with the manuscript.

